# Brain Health Loss Mediates the Effect of Infarct Volume on Functional Outcome in Ischemic Stroke

**DOI:** 10.1101/2025.09.26.25335406

**Authors:** Erik Lindgren, Luca Angeleri, Kenda Alhadid, Christina Jern, Arne G Lindgren, Jane Maguire, Robert W. Regenhardt, Natalia S. Rost, Markus D. Schirmer, the MRI-GENIE and GISCOME Investigators

**Affiliations:** J. Philip Kistler Stroke Research Center, Massachusetts General Hospital, Harvard Medical School, Boston, MA, USA; Department of Clinical Neuroscience, Institute of Neuroscience and Physiology, Sahlgrenska Academy, University of Gothenburg; Department of Neurology, Sahlgrenska University Hospital, Region Västra Götaland, Gothenburg, Sweden; Department of Laboratory Medicine, Institute of Biomedicine, Sahlgrenska Academy, University of Gothenburg; Department of Clinical Genetics and Genomics, Sahlgrenska University Hospital, Region Västra Götaland, Gothenburg, Sweden; Department of Neurology, Skåne University Hospital; Department of Clinical Sciences Lund, Neurology, Lund University, Lund, Sweden; University of Technology Sydney, Sydney, Australia

**Author notes:** **Corresponding Author:** Erik Lindgren, MD, PhD, Department of Neurology, Sahlgrenska University Hospital, and Department of Clinical Neuroscience, Institute of Neuroscience and Physiology, Sahlgrenska Academy at University of Gothenburg, Blå Stråket 7, 3rd floor, 413 45, Gothenburg, Sweden.

**Keywords:** brain health, stroke, acute ischemic stroke, outcome, imaging, mediation

## Abstract

**Importance:** Brain health may facilitate resilience to detrimental consequences from neurological diseases. Infarct volume is associated with poor functional outcome after acute ischemic stroke (AIS), but potential mediating effects through stroke-related brain health loss have not been investigated.

**Objective:** To determine whether stroke-related brain health loss, quantified by change in MRI derived effective Reserve (eR), mediates the effect of acute infarct volume on functional outcome after AIS.

**Design:** Observational multicenter cohort study.

**Setting:** We analyzed data from the GASROS (n=488) and MRI-GENIE (n=560) cohorts, collected 2003-2011.

**Participants:** Adult patients consecutively diagnosed with AIS, with available admission MRI.

**Exposure:** At admission, white matter hyperintensity (WMH) and normal-appearing brain volumes were assessed on T2-FLAIR, and acute infarct volume on diffusion weighted imaging. WMH was normalized by brain volume, creating WMH load. We quantified brain health using eR, a latent variable incorporating age, WMH load, and normal-appearing brain volume. ΔeR reflected the change in eR when acute infarct volume was included, representing stroke-related brain health decline. Mediation analysis was used to determine if ΔeR mediates the effect of infarct volume on functional outcome (modified Rankin Scale [mRS] at 90 days).

**Main Outcome Measure:** Proportion of mediating effect.

**Results:** We included 1,048 patients (median age 67y, 38% females). At baseline, median NIHSS score was 3 (IQR 1-7), median infarct volume 3.1mL (IQR 0.9-15.5). At 90 days, median mRS score was 1 (IQR 1-3) and 51 (5%) patients had died. In mediation analysis, ΔeR significantly mediated 36% (95% CI 16-56%) of the total effect of infarct volume on functional outcome (direct effect (ß=0.15 [95% CI 0.09-0.22], p<0.001; indirect effect mediated through ΔeR: ß=0.09 [95% CI 0.04 to 0.14], p=0.001). In subgroup-analyses, the mediative effect was apparent among female but not male, and among patients aged >67y but not ≤67y.

**Conclusions and Relevance:** Stroke-related structural brain health loss mediates about one third of the effect of acute infarct volume on functional outcome after ischemic stroke, with important sex and age differences. Brain health significantly influences outcome and recovery potential, and may be considered a key biomarker when modeling outcome after AIS.

**Key points:** *Question:* Does brain health loss associated with acute ischemic stroke mediate the relationship between acute infarct volume and functional outcome?

*Findings:* In this observational multicenter cohort study of 1,048 patients, mediation analysis suggests that reduction of the brain health MRI marker effective Reserve mediates 36% (95% CI 16-56%) of the total effect of acute infarct volume on functional outcome. The proportion of mediative effect was more pronounced among female compared to male and in older compared to younger patients.

*Meaning:* Brain health loss mediates one third of the effect of acute infarct volume on functional outcome after ischemic stroke.

## Introduction

Stroke is a leading contributor to long-term disability and death, with 70 million ischemic stroke survivors worldwide.^1-4^ Debilitating consequences affect patients, their relatives, and societies, with costs for functional deficits exceeding $50 billion annually in the US alone.^3^ It is essential to comprehend mechanisms which facilitate recovery and functional independence after stroke.^5^ Brain health is a progressing concept encompassing both the absence of disease and the brain’s optimal functioning during a lifespan.^6,7^ The protective mechanisms of brain health may include resilience to endure or even avoid neurological diseases.^8^ Although brain health is associated with retained cognitive function while aging and in neurodegenerative diseases,^3^ its role in functional recovery after acute ischemic stroke (AIS) remains poorly understood.

Prominent international organizations’ calls to action to optimize brain health have led to increased societal and professional awareness.^6,9^ However, the inherent complexity of brain health presents substantial hurdles to establish measures that are both assessable in a research setting and implementable in clinical practice.^6,7,9^ To meet this need, a quantifiable magnetic resonance imaging (MRI) marker of brain health called *effective Reserve* (eR) has been developed, and associated with functional outcome after AIS.^10,11^ An individual’s eR can be estimated by using automated pipelines calculating volumetrics based on routinely acquired MRI.^10,11^

While infarct volume is a strong predictor for poor functional outcome after AIS,^12^ its role in affecting brain health has not been thoroughly investigated. We hypothesize that acute stroke-related brain health loss, estimated by change in eR, partially mediates the negative effects of infarct volume on functional outcome after stroke.

## Methods

### Standard protocol approval, registration and patient consent

The study was conducted in accordance with the 1964 Declaration of Helsinki and its amendments. Each center received permission from local ethics committees to collect observational data as required under applicable national laws.

The study is reported following international guidelines for reporting observational studies, Strengthening the Reporting of Observational Studies in Epidemiology (STROBE) statement^13^ and the Sex and Gender Equity in Research (SAGER) guidelines.^14^

### Participants

We analyzed pooled data from the Genes Affecting Stroke Risk and Outcomes Study (GASROS), and MRI-GENetics Interface Exploration (MRI-GENIE) cohorts. Details of both GASROS and MRI-GENIE have been described previously.^15,16^ GASROS is a single center cohort including adult patients (age ≥ 18 years) presenting with symptoms and signs of AIS. MRI-GENIE is an international multicenter study of hospital-based cohorts covering adult patients diagnosed with AIS. The number of participants analyzed per center is shown in supplement eTable 1. Both cohorts’ inclusion periods spanned from 2003 to 2011. Patients with available admission MRI including T2-Fluid-attenuated inversion recovery (FLAIR) sequences and acute infarct lesion on diffusion weighted imaging (DWI) were eligible for inclusion.

### Neuroimaging data curation

Neuroimaging was obtained at acute hospital admission as part of routine care using clinical MRI AIS protocols with median time to scan of 2 days (IQR 1-4 days). All images were either obtained on 1T, 1.5T or 3T scanners (General Electric Medical Systems, Philips Medical Systems, Siemens, Toshiba, Marconi Medical Systems, Picker International Inc.). Specific MRI features included for each participant: T2-FLAIR imaging (Repetition time 5000 ms, minimum Echo time of 62 to 116 ms, Inversion time 2200 ms, Field-of-view 220-240 mm) and DWI (single-shot echo-planar imaging; 1-5 B0 volumes, 6-30 diffusion directions with b=1000 s/mm^2^, 1-3 averaged volumes). We used automated deep learning enabled pipelines to assess white matter hyperintensity volume (WMHv)^17^ and brain volume (combined gray matter and white matter parenchymal volumes) based on FLAIR,^16,18^ and acute infarct volume based on DWI sequences.^19^ Normal appearing brain volumes were calculated subtracting WMHv from brain volume for each patient. The automated pipelines have been developed specifically for analysis on routinely acquired MRI sequences of patients with AIS.^16-19^ Each sequence and automatically segmented mask (brain volume, WMH volume and stroke lesion volume) was manually quality checked by visual inspection, excluding cases with severe motion artifacts and/or apparent segmentation failure. We normalized WMHv and lesion volume by individual brain volumes, creating WMH load and lesion load for each patient.

### Data points and definitions

Data on clinical characteristics, demographics, and medical history were collected on hospital admission and at follow-up. Ischemic stroke etiological subtypes were categorized according to the Trial of Org 10172 in Acute Stroke Treatment (TOAST) classification.^20^ Functional outcome and mortality was assessed approximately 90 days after stroke onset using the modified Rankin Scale (mRS) based on in-person visits or interview by telephone.^21^ Functional outcome was analyzed among survivors as an ordinal variable with mRS scores ranging from 0-5 (0 indicating no symptoms, 5 indicating severe disability) and mortality was analyzed separately as a dichotomized variable in sub-analysis.

### Brain health MRI marker and brain health change

We defined eR as a latent variable in structural equation modeling (SEM) based on the measured variables: age, WMH load (logit transformed for normal distribution) and normal-appearing brain volume.^10,11^ We defined acute stroke-related structural brain health loss as the change in eR (ΔeR) pre- and post-stroke as follows: Assuming the infarct volume as healthy tissue, estimated eR without the lesion represented the pre-stroke state vs. estimated eR with the lesion volume as infarct included in the post-stroke state. The pathway coefficients to estimate eR from the observed variables were calculated using bootstrapping (100 samples), using patients from MRI-GENIE cohort with available FLAIR imaging and outcome data. These coefficients were used to estimate eR for each patient across cohorts.

### Primary investigated measure

The primary investigated measure was the proportion of the mediated effect through ΔeR of the total effect of acute infarct volume on functional outcome at 90 days.

### Statistical analysis

We report continuous data as medians with interquartile ranges (IQR, 25^th^ to 75^th^ percentile), and dichotomous data as proportions of investigated patients. For comparative analysis, the significance level was set to p<0.05, using Mann-Whitney U test for continuous data and Chi-square test for dichotomous data. We assessed normal distributions using Shapiro-Wilk test and applied logit transformation of variables when applicable.

In accordance with guidelines from the Health Insurance Portability and Accountability Act (HIPAA), participants with age 90 years or older were aggregated into a single age category. Patients with incomplete data of any of the key variables (age, sex, WMH volume, infarct volume, brain volume) were excluded. Missing data are reported for each of the investigated variables separately.

The hypothesized causal model where ΔeR mediates the effect of infarct volume on functional outcome at 90 days is illustrated in Figure 2. Previous research established the association between acute infarct volume and functional outcome after stroke.^22,23^ Using mediation analysis in SEM, we defined ΔeR as the mediator and calculated beta coefficients for the indirect pathway (infarct volume-ΔeR-functional outcome), direct pathway (infarct volume-functional outcome), and total effect (indirect plus direct pathways). If the total and indirect effects were both significant (p<0.05), we also calculated the proportion of the mediated effect (ratio of indirect effect to total effect). We used bootstrapping (100 iterations) to calculate standardized pathway coefficients with standardized 95% confidence intervals (CI) and p-values. All analyses were performed using R statistic programming version 4.4.1 (R Foundation for Statistical Computing)^24^ and for latent variable and mediation analysis, the statistical package Lavaan.^25^

### Sub-Analyses

In a first sub-analysis, we used the same methodology to calculate pathway coefficients described in Figure 2, stratified by sex. The magnitude of the coefficients and proportions of mediated effect were illustrated in a forest plot. Secondly, we repeated the same analysis categorizing patients into older and younger dichotomized by the median age. Thirdly, we calculated pathway coefficients using mortality within 90 days as outcome variable. Fourthly, we repeated the main analysis in GASROS and MRI-GENIE cohorts separately.

### Data availability statement

Data will be made available for the purpose of reproducing the results, upon reasonable request to the corresponding author, at the discretion of local Institutional Review Boards.

## Results

In total 1,210 patients with AIS, acute MRI including T2-FLAIR and DWI sequences and outcome data at 90 days were eligible for inclusion. We excluded 162 patients (13.4%) due to insufficient imaging quality or incomplete data of key variables (Figure 1).

**Figure 1.**
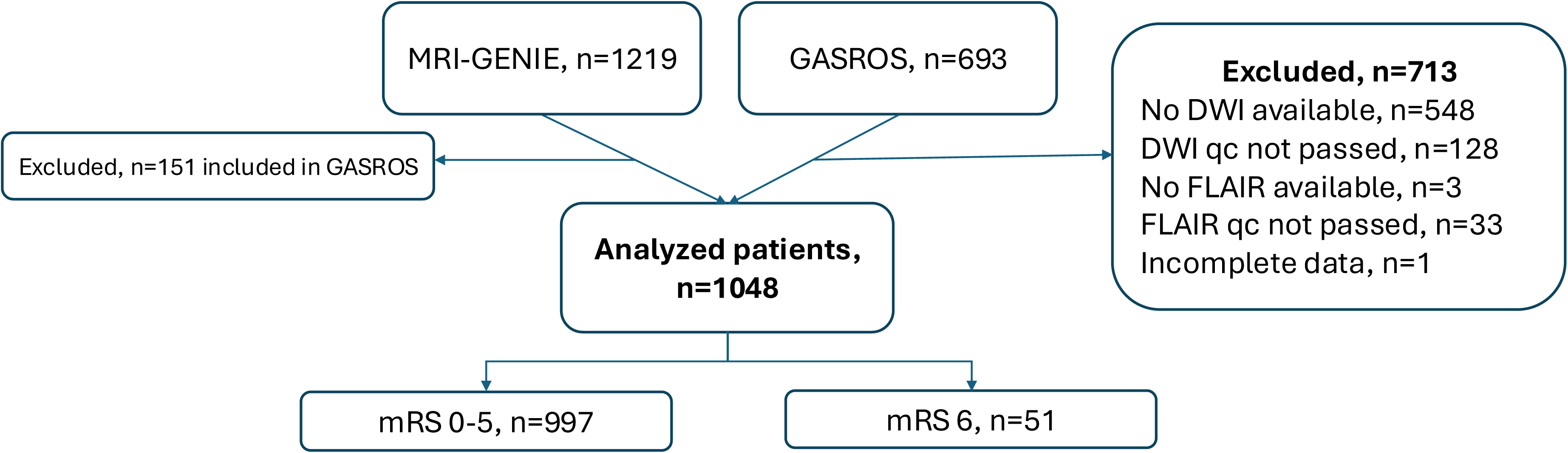
Selection of Patients with Acute Ischemic Stroke Included in the Mediation Analysis. *DWI – diffusion weighted imaging; FLAIR – fluid attenuated inversion recovery; qc – quality check; MRI-GENIE - MRI-GENetics Interface Exploration study; GASROS - the Genes Affecting Stroke Risk and Outcomes Study*

We analyzed 1,048 patients (n=488 from GASROS and n=560 from MRI-GENIE) with a median age of 67 years (IQR 56-77), of which 401 (38%) were female. Baseline characteristics and outcomes for the pooled cohort and stratified by sex are depicted in Table 1. At baseline, median NIHSS score was 3 (IQR 1-7), median brain volume 1300 mL (IQR 1190-1400) and median acute infarct volume 3.1 mL (IQR 0.9-15.5). At 90 days, median mRS score was 1 (IQR 1-3), and 51 (5%) patients had died. At onset, female patients were older, had relatively smaller calculated brain volumes, more frequent history of hypertension and less frequent history of smoking as compared to males. Acute infarct volume, infarct load, WMH load, NIHSS score, and mortality rates did not differ between sexes, but females had higher median mRS score at 90 days.

**Table 1.** Baseline Characteristics and Outcomes.

| Variable | No. (%) |  |  | P-value |
| --- | --- | --- | --- | --- |
|  | Total (n=1048) | Female (N=401) | Male (N=647) |  |
| Demographics |  |  |  |  |
| Age, median (IQR), y | 67.0 (56.0-76.6) | 70.2 (55.9-79.6) | 66.0 (56.0-74.0) | <0.001 |
| Female | 401 (38.3) | NA | NA |  |
| Cardiovascular Risk Factors |  |  |  |  |
| Type 2 Diabetes Mellitus | 220/1042 (21.0) | 77/399 (19.2) | 143/643 (22.1) | 0.292 |
| Hypertension | 670/1048 (63.9) | 274/400 (68.3) | 396/646 (61.2) | 0.022 |
| Smoking | 620/1031 (59.2) | 194/392 (48.4) | 426/639 (65.8) | <0.001 |
| Atrial Fibrillation | 386/1038 (36.8) | 146/397 (36.4) | 240/641 (37.1) | 0.881 |
| Clinical presentation |  |  |  |  |
| NIHSS, median (IQR) | 3 (1-7)<sup>a</sup> | 3 (1-8)<sup>b</sup> | 3 (1-6)<sup>c</sup> | 0.068 |
| TOAST classification |  |  |  |  |
| Large-artery atherosclerosis | 176 (16.8) | 52 (13.0) | 124 (19.2) | 0.024 |
| Cardioembolism | 317 (30.2) | 122 (30.4) | 195 (30.1) |  |
| Small Vessel Occlusion | 144 (13.7) | 52 (13.0) | 92 (14.2) |  |
| Stroke of Other Determined Etiology | 135 (12.9) | 53 (13.2) | 82 (12.7) |  |
| Stroke of Undetermined Etiology | 183 (17.5) | 85 (21.2) | 98 (15.1) |  |
| Missing | 93 (8.9) | 37 (9.2) | 56 (8.7) |  |
| Imaging volumetrics, median (IQR) |  |  |  |  |
| Brain Volume (mL) | 1300 (1190-1400) | 1210 (1120-1290) | 1360 (1260-1450) | <0.001 |
| White Matter Hyperintensity Volume (mL) | 6.18 (2.37-16.2) | 5.98 (2.27-16.1) | 6.47 (2.46-16.2) | 0.575 |
| White Matter Hyperintensity Load (%) | 0.50 (0.18-1.32) | 0.51 (0.18-1.42) | 0.48 (0.17-1.26) | 0.411 |
| Acute Infarct Volume (mL) | 3.13 (0.90-15.5) | 3.24 (0.97-16.5) | 3.08 (0.89-15.2) | 0.749 |
| Acute Infarct Load (%) | 0.25 (0.07-1.24) | 0.27 (0.08-1.40) | 0.24 (0.06-1.16) | 0.194 |
| Normal appearing brain volume (pre, IQR) | 1290 (1180-1390) | 1200 (1100-1270) | 1350 (1250-1440) | <0.001 |
| Normal appearing brain volume (post, IQR) | 1270 (1170-1380) | 1180 (1090-1260) | 1340 (1230-1430) | <0.001 |
| Outcome at 90 days |  |  |  |  |
| mRS score, median (IQR) | 1 (1-3) | 2 (1-3) | 1 (1-2) | <0.001 |
| 0 | 225 (21.5) | 64 (16.0) | 161 (24.9) |  |
| 1 | 352 (33.6) | 120 (29.9) | 232 (35.9) |  |
| 2 | 185 (17.7) | 72 (18.0) | 113 (17.5) |  |
| 3 | 135 (12.9) | 70 (17.5) | 65 (10.0) |  |
| 4 | 88 (8.4) | 41 (10.2) | 47 (7.3) |  |
| 5 | 51 (4.9) | 8 (2.0) | 4 (0.6) |  |
| Mortality | 51/1048 (4.9) | 26 (6.5) | 25 (3.9) | 0.077 |

<sup>a</sup> 12 patients missing. <sup>b</sup> 5 patients missing. <sup>c</sup> 7 patients missing. IQR – Inter Quartile Range, mL – milliliter, mRS – modified Rankin Scale

### Mediation analysis

The main mediation analysis was performed in 997 patients with mRS 0-5 at 90 days and the pathway coefficients are summarized in Table 2. Acute infarct volume was independently associated with worse functional outcome (total effect ß=0.24 [95% CI 0.20 - 0.30], p<0.001), of which 36% (95% CI 16.3-56.4%) was mediated by ΔeR (indirect effect ß=0.09 [95% CI 0.04 - 0.14], p=0.001).

**Table 2.** Change in Effective Reserve Mediates the Total Effect of Acute Lesion Volume and Functional Outcome at 90 days After Acute Ischemic Stroke.

| Functional Outcome | All patients (n=997) |  |  | Female (n=375) |  |  | Male (n=622) |  |  |
| --- | --- | --- | --- | --- | --- | --- | --- | --- | --- |
| | $\beta$ | (95% CI) | p-value | $\beta$ | (95% CI) | p-value | $\beta$ | (95% CI) | p-value |
| **a** | 0.692 | (0.620 – 0.759) | 0.000 | 0.687 | (0.560 - 0.833) | 0.000 | 0.696 | (0.648 – 0.779) | 0.000 |
| **b** | 0.124 | (0.054 – 0.198) | 0.001 | 0.249 | (0.134 - 0.412) | 0.000 | 0.056 | (-0.060 to 0.167) | 0.314 |
| **Direct effect** | 0.152 | (0.089 – 0.220) | 0.000 | 0.053 | (-0.060 to 0.146) | 0.296 | 0.216 | (0.107- 0.294) | 0.000 |
| **Indirect effect (a\*b)** | 0.086 | (0.040 – 0.141) | 0.001 | 0.171 | (0.092 – 0.335) | 0.001 | 0.039 | (-0.043 to 0.123) | 0.319 |
| **Total effect** | 0.238 | (0.196 – 0.295) | 0.000 | 0.224 | (0.139 – 0.349) | 0.000 | 0.255 | (0.201 – 0.318) | 0.000 |
| **Proportion of mediated effect (indirect/direct)** | 36% | (16.3- 56.4%) | NA | 76.3 % | (41.5-100%) | NA | NA | NA | NA |

### Sex differences

Stratifying by sex, the indirect effect via ΔeR was apparent among females (indirect effect ß=0.17, [95% CI 0.09-0.34], p=0.001, proportion of mediated effect 76.3%, [95% CI 41.5 – 100%]), but not among males (indirect effect ß=0.04 [95% CI -0.04 to 0.12], p=0.319). Comparisons of the mean estimates and 95% CIs are illustrated graphically in Figure 3.

**Figure 2.**
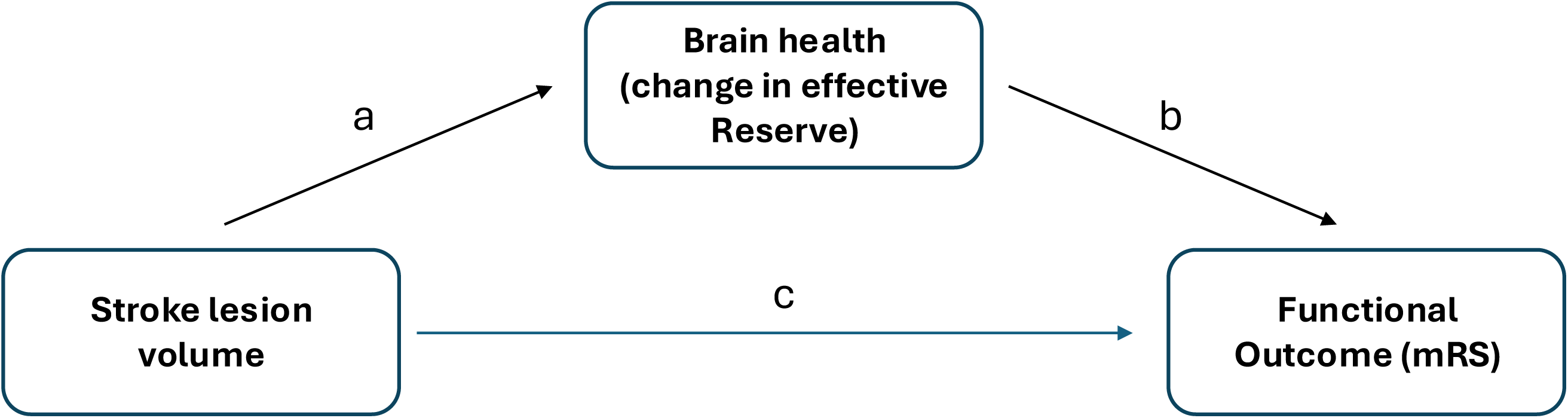
Model of the Hypothetical Causal Pathways in Patients with Acute Ischemic Stroke. *We defined brain health loss, quantified using change in the effective Reserve caused by stroke, as a potential mediator influencing the effect of stroke lesion volume on functional outcome. The direct effect is defined as pathway c, and the indirect effect as pathways a x b. Outcomes were assessed at 90 days after onset of acute ischemic stroke. Functional outcome was analyzed among survivors as an ordinal variable with modified Rankin Scale scores ranging from 0-5 (0 indicating no symptoms, 5 indicating severe disability). In a sub-analysis, mortality as outcome was analyzed as a dichotomized variable*.

**Figure 3.**
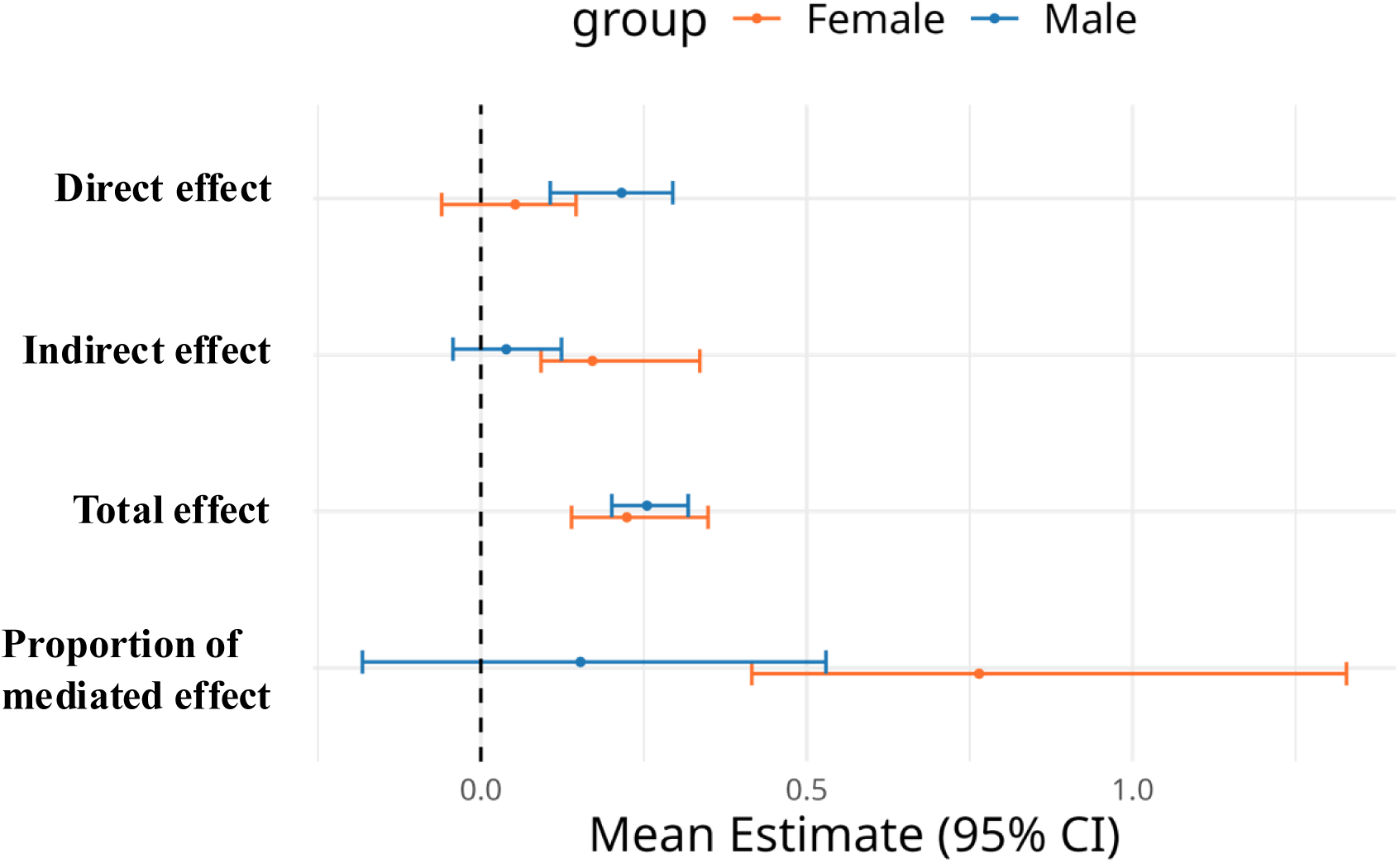
Sex Differences in Mediation Pathway Mean Estimates. *The hypothetical causal model where effective Reserve loss mediates the effect of infarct volume on functional outcome at 90 days is illustrated in Figure 2. Bootstrapped mean estimates of each pathway and 95% confidence intervals. Direct effect, indirect effect, total effect and proportion of mediated effect differed significantly between females and males according to Mann-Whitney U tests (p<0.001)*.

### Age differences

The mediative effect of ΔeR was apparent in the older patient group (>67y, n=480 patients, indirect effect ß=0.16 [95% CI 0.08 – 0.26, p<0.001], proportion of mediated effect 75.4% [40.6-100%]) but not in the younger patient group (≤67y, n=517, indirect effect ß=0.016 [95% CI -0.06 to 0.10, p=0.709]). Pathway coefficients are further described in eTable 2 and mean estimates with 95% CIs are illustrated graphically in eFigure 1.

### Mediative Effect on Mortality

We included 1,048 patients in a sub analysis to evaluate the same model as described in Figure 2, using mortality as outcome. Acute infarct volume was independently associated with mortality at 90 days (total effect ß=0.167 [95% CI 0.076-0.264], p<0.001) but we observed no mediative effect through ΔeR (eTable 3).

Comparisons between cohorts are available in the eResults and eTable 4-5 in the Supplement.

## Discussion

In this observational, multicenter cohort study of 1,048 patients with ischemic stroke, acute stroke-related brain health loss, quantified using the MRI marker eR, significantly mediated 36% of the total effect of stroke lesion volume on functional outcome. The mediative effects were more pronounced among females and in older patients. Our results support biomarkers of structural brain health to be considered key variables in future studies investigating outcome and recovery after AIS.

Lesion volume has consistently been reported to be an independent predictor of poor outcome after ischemic stroke,^12^ but potential mediative effects of acute stroke-related brain health loss have not previously been explored. Predictive models considering phenotypical factors alone (e.g. age, cardiovascular risk factors, and comorbidities) cannot fully explain the variability in individuals’ capacities to recover after ischemic stroke. Our results partially bridge that knowledge gap, proposing eR as a meaningful biomarker for resilience to poor functional outcome after AIS. Potential protective mechanisms for individuals with preserved brain health include enhanced neuroplasticity, reportedly associated with intact neurons and white matter integrity facilitating functional recovery.^8^ Future studies may expand on MRI brain health markers to incorporate also lesion location and brain health spatial patterns.^26-29^

Brain health has been endorsed as a global research focus goal.^6,7,9^ Robust, reproducible brain health markers, taking the inherent complexity of brain health into account while effectively balancing with automatization and simplicity for practical clinical use are warranted. While several measures capturing brain health exist, few direct comparisons on their predictive performances after AIS have been conducted.^30^ Investigating structural brain health, most studies have focused on isolated measures of small vessel disease,^31^ WMH,^32,33^ and regional or global atrophy.^34^ The additive value of including composite or comprehensive measures of brain health, or conversely of brain frailty have been emphasized, but generally rely on manual assessment or high quality imaging, often not obtainable in an acute stroke setting.^10,34-38^

A recent comparative study suggests eR as a more meaningful biomarker to predict functional outcome as compared to the measured variables age, WMH, and normal appearing brain volume separately, compared to clinical variables alone and compared to other available structural measures of brain health.^30^ In this study, we advance the concept of eR to include normal-appearing brain volume rather than brain volume, and further introduce a model approximating acute stroke-related brain health loss.^11^ In all, our results are in line with the growing body of evidence showing brain health markers to be linked to outcome after stroke. For example, in 1105 patients randomized to either potentially neuroprotective treatment or best medical care after EVT, neuroimaging frailty mediated 85% of the total effect of age on 90-day functional outcome.^39^ In another study, higher radiomics derived brain age has been reported associated with worse outcomes after AIS.^36,40^ Manually assessed imaging brain frailty is associated with higher NIHSS scores at admission and worse recovery trajectories after EVT.^38^ Our results additionally establish a hypothesis that infarcts partially influence functional outcome after stroke through acute brain health loss.

We acknowledge that our estimation of brain health loss may be influenced by early morphological changes or infarct visibility on FLAIR, since the median scan time was 2 days post-admission, and that time from index stroke to admission was not known. However, such confounding would probably lead to an underestimation of eR change, thereby reducing the chance of detecting an underlying mediating effect. Further, eR has not yet been shown to capture cognitive brain health and does not directly account for regional brain health differences, biological age, proteomics, genetics, social interactions, or cognition. Future measures of brain health may be expanded to also include these areas, but these goals must be balanced to account for increasing complexity.

A key novel finding is that the mediative effect of acute brain health loss on functional outcome was more pronounced among females compared to males. Sex differences in stroke are apparent in epidemiologic patterns, presentation, etiologies, treatments, and the burden of disability after stroke is disproportionally skewed toward females. For example, females are on average 4-6 years older than men at stroke onset,^41^ females have a higher lifetime risk of stroke,^42^ and the negative impact of cardiovascular risk factors among females entails higher risks for stroke as compared to males. Females are exposed to hormonal changes of estradiol as well as pre-/menstrual, pregnancy and birth related conditions which reportedly affect the risk of stroke.^43-46^

Analogously, it is not unlikely that the protective importance of brain health would also differ between sexes. In our study, females had worse functional outcome at 90 days although sex differences in normalized lesion volume or NIHSS score at onset were not observed. Higher age adjusted WMH load has been reported among females, but was not reproduced in our study.^33^ The pipeline to quantify WMH has shown comparable results as manual segmentation and using other validated methods for WMH assessment are not likely to substantially influence the results.^17^ Comparatively lower eR among females attributable to higher age at onset and relatively lower brain volumes may lead to worse outcomes, but do not directly explain the observed differences in the mediative effect. Hypothetically, females may reach the threshold for which a diminished reserve presents as clinical consequences earlier than males.^8^ Recent longitudinal imaging studies have shown noteworthy structural changes with reduced white matter integrity and regional grey matter loss during the course of pregnancy, some of which seem long-lasting suggesting considerable impact of female specific hormonal changes during a lifespan.^47^ The biological and psychosocial mechanisms underlying sex differences in the importance of brain health and stroke should be targeted in future research.

Our results further imply important age differences, with a stronger mediative effect of brain health among older patients as compared to younger patients. These findings resonate with Statz theory that the brain posits a reserve capacity, and once the brain reserve capacity is depleted past a fixed critical threshold, clinical deficits emerge.^8^ As the reserve declines while aging, older patients have relatively lower reserve and thereby also slimmer margins when affected by a sudden vascular event. Hence, what is left matters more for older patients.

Several additional limitations in our study warrant comment. First, while the mediative effect of brain health loss was apparent in the pooled cohort, we could not observe significant mediative effects when analyzing the cohorts separately, probably due to insufficient number of patients in relation to the effect size (eResults, eTable 4 and eTable 5). Second, low- and middle-income countries were not represented and the generalizability to regions not represented in the included cohorts is limited. Third, we did not have the statistical power to perform subgroup analyses stratified by stroke subtype.^48^ Fourth, our cohorts included patients between 2003-2011 with mostly mild to moderate stroke severity, with limited data on both intravenous thrombolysis and endovascular treatment, and may not be fully representative of patients treated in today’s stroke care settings.

To conclude, we report salient insights on the underlying mechanisms through which infarct volume and functional outcome relate. Our results add to recent evidence of brain health being associated with outcomes after stroke,^11,36^ that brain health acts as an important mediator in the casual pathway connecting stroke lesion volume to functional outcome. We show that acute stroke-related brain health loss mediates one third of the effect of lesion volume on functional outcome after ischemic stroke, emphasizing the importance of brain health for recovery after stroke. Our results support that measures of structural brain health, while taking sex and age into account, may be considered key variables in future studies investigating outcome and recovery after AIS and reinforce the necessity to identify and implement broad strategies to optimize and improve brain health for all.

## Acknowledgments

The authors thank participating patients, their families and investigators of the MRI-GENIE and GISCOME collaborations.

