## Supplement for "Brain Health Loss Mediates the Effect of Infarct Volume on Functional Outcome in Ischemic Stroke"

**eResults**

*Comparing GASROS vs MRIGENIE cohorts*

Baseline characteristics are compared between the included cohorts in the supplement eTable 4. Patients included from GASROS had lower NIHSS score at baseline, more often history of smoking, higher proportion of patients with atrial fibrillation, larger brain volumes and smaller acute infarct volumes as compared to MRI-GENIE. Functional outcome at 90 days was more favorable, but all-cause mortality rates were higher in GASROS as compared to MRI-GENIE. The acute infarct volume was independently associated with poor functional outcome at 90 days, but we found no significant mediation effect by ΔeR in either of the cohorts when analyzed separately (pathway coefficients presented in the supplement eTable 5).

**eFigure 1. Age Differences in Mediation Pathway Mean Estimates**

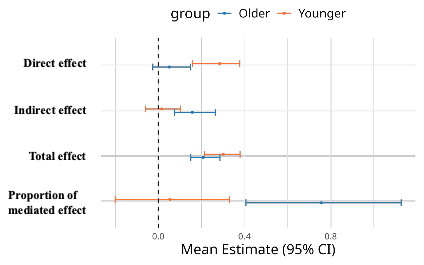

*The hypothetical causal model where effective Reserve mediates the effect of infarct volume on functional outcome at 90 days is illustrated in Figure 2. eFigure 1 describes bootstrapped mean estimates of each pathway and 95% confidence intervals stratified by median age 67 years. Direct effect, indirect effect, total effect and proportion of mediated effect differed significantly between relatively younger and older patients, according to Mann-Whitney U tests (p<0.001).*

**eFigure 2. Selection of Patients with Acute Ischemic Stroke Included in the Mediation Analyses by Cohort**

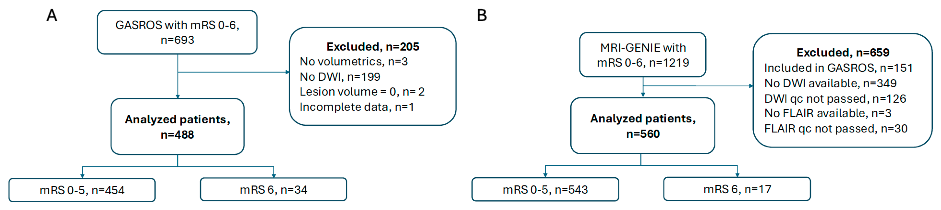

**eTable 1. Participating Centers and Included Patients**

| **Country** | **Center** | **Included Patients** |
| --- | --- | --- |
| ***MRI-GENIE*** | | ***N=560*** |
| Belgium | University Hospitals, Leuven | 299 (53%) |
| Sweden | Skåne University Hospital Lund, Lund (Lund Stroke Register) | 74 (13%) |
| Sweden | Sahlgrenska University Hospital, Gothenburg (SAHLSIS) | 50 (9%) |
| Spain | IMIM-Hospital del Mar, Barcelona, (BASICMAR) | 65 (12%) |
| USA | University of Cincinnati, OH (GCNKSS) | 72 (13%) |
| ***GASROS*** | | ***N=488*** |
| USA | Massachusetts General Hospital, MA | 488 (100%) |

*MRI-GENIE - MRI-GENetics Interface Exploration study; GASROS - the Genes Affecting Stroke Risk and Outcomes Study*

**eTable 2. Pathway Coefficients Assessing Mediative Effects of Effective Reserve on the Relationship Between Lesion Volume and Functional Outcome at 90 days, Stratified by Median Age**

| Functional Outcome | > 67 years (n=480) | | | ≤ 67 years (n=517) | | |
| --- | --- | --- | --- | --- | --- | --- |
|  | **ß** | **(95% CI)** | **p-value** | **ß** | **(95% CI)** | **p-value** |
| **a** | 0.681 | (0.608 – 0.817) | 0.000 | 0.698 | (0.599 – 0.791) | 0.000 |
| **b** | 0.231 | (0.110 – 0.359) | 0.000 | 0.023 | (-0.080 to 0.138) | 0.711 |
| **Direct effect** | 0.051 | (-0.024 to 0.151) | 0.242 | 0.285 | (0.160 - 0.378) | 0.000 |
| **Indirect effect (a*b)** | 0.157 | (0.075 – 0.264) | 0.000 | 0.016 | (-0.058 to 0.103) | 0.709 |
| **Total effect** | 0.209 | (0.151 – 0.286) | 0.000 | 0.301 | (0.215 – 0.380) | 0.000 |
| **Proportion of mediated effect (indirect/direct)** | 75.4% | (40.6% – 100%) | NA | NA | NA | NA |

**eTable 3. Pathway Coefficients Assessing Mediative Effects of Effective Reserve on the Relationship Between Lesion Volume and Mortality at 90 days**

| Mortality | All patients (n=1,048) | | |
| --- | --- | --- | --- |
|  | **ß** | **(95% CI)** | **p-value** |
| **a** | 0.696 | (0.637 – 0.769) | 0.000 |
| **b** | 0.069 | (-0.083 to 0.386) | 0.516 |
| **Direct effect (c)** | 0.118 | (-0.160 to 0.246) | 0.183 |
| **Indirect effect (a*b)** | 0.048 | (-0.060 to 0.251) | 0.516 |
| **Total effect** | 0.167 | (0.076 – 0.264) | 0.000 |
| **Proportion of mediated effect (indirect/direct)** | NA | NA | NA |

**eTable 4. Baseline Characteristics GASROS and MRI-GENIE Cohorts**

| Variable | No. (%) | | P-value | |
| --- | --- | --- | --- | --- |
|  | **GASROS** (N=488) | **MRI-GENIE**  (N=560) |  |  |
| *Demographics* | | | | |
| Age, median (IQR), y | 65.8 (55.0-76.3) | 68.0 (57.0-77.0) | | 0.131 |
| *Cardiovascular Risk Factors* | | | | |
| Type 2 Diabetes Mellitus | 96/488 (19.7) | 124/554 (22.1) | | 0.32 |
| Hypertension | 300/488 (61.5) | 370/558 (66.1) | | 0.119 |
| Smoking | 319/485 (65.4) | 301/546 (53.8) | | **<0.001** |
| Atrial Fibrillation | 283/488 (58.0) | 103/550 (18.4) | | **<0.001** |
| *Clinical presentation* | | | | |
| NIHSS, median (IQR) 5 and 7 missing | 3 (1-6)* | 4 (2-7) | | **<0.001** |
| TOAST classification | | | | |
| Large-artery atherosclerosis | 87 (17.8) | 89 (15.9) | | **<0.001** |
| Cardioembolism | 175 (35.9) | 142 (25.4) | |  |
| Small Vessel Occlusion | 58 (11.9) | 86 (15.4) | |  |
| Stroke of Other Determined Etiology | 113 (23.2) | 22 (3.9) | |  |
| Stroke of Undetermined Etiology | 36 (7.4) | 147 (26.3) | |  |
| Missing | 19 (3.9) | 74 (13.2) | |  |
| *Imaging volumetrics, median (IQR)* | | | | |
| Brain Volume (mL) | 1320 (1210-1420) | 1280 (1180-1390) | | **<0.001** |
| White Matter Hyperintensity Volume (mL) | 6.13 (2.10-16.8) | 6.23 (2.57-15.7) | | 0.527 |
| White Matter Hyperintensity Load (%) | 0.47 (0.16-1.3) | 0.52 (0.20-1.29) | | 0.360 |
| Acute Infarct Volume (mL) | 2.26 (0.553-13.4) | 4.21 (1.32-17.0) | | **<0.001** |
| Acute Infarct Load (%) | 0.17 (0.05-1.03) | 0.33 (0.10-1.39) | | **<0.001** |
| Normal appearing brain volume (pre, IQR) | 1310 (1190-1420) | 1260 (1160-1370) | | **<0.001** |
| Normal appearing brain volume (post, IQR) | 1300 (1180-1410) | 1250 (1150-1360) | | **<0.001** |
| *Outcome at 90 days* | | | | |
| mRS score, median (IQR) | 1 (0-3) | 1 (1-3) | | **<0.001** |
| Mortality | 34 (7.0) | 17 (3.0) | | **0.005** |

**eTable 5. Pathway Coefficients Assessing Mediative Effects of Effective Reserve on the Relationship Between Lesion Volume and Functional Outcome at 90 days, Stratified by Investigated Cohorts**

| Functional outcome | GASROS (n=454) | | | MRIGENIE (n=543) | | |
| --- | --- | --- | --- | --- | --- | --- |
|  | **ß** | **(95% CI)** | **p-value** | **ß** | **(95% CI)** | **p-value** |
| **a** | 0.692 | (0.633 – 0.780) | 0.000 | 0.705 | (0.622 – 0.824) | 0.000 |
| **b** | 0.070 | (-0.048 to 0.215) | 0.229 | 0.110 | (-0.017 to 0.225) | 0.055 |
| **Direct effect** | 0.223 | (0.122 – 0.312) | 0.000 | 0.124 | (0.026 – 0.228) | 0.014 |
| **Indirect effect (a*b)** | 0.048 | (-0.034 to 0.150) | 0.234 | 0.078 | (-0.012 to 0.163) | 0.059 |
| **Total effect** | 0.271 | (0.212 – 0.318) | 0.000 | 0.201 | (0.134 – 0.304) | 0.000 |
| **Proportion of mediated effect (indirect/direct)** | NA | NA | NA | NA | NA | NA |

**eTable 6. MRI-GENIE Study Group Members**

| **Name** | **Location** | **Role** | **Contribution** |
| --- | --- | --- | --- |
| Markus D. Schirmer, PhD | J. Philip Kistler Stroke Research Center, Massachusetts General Hospital, Harvard Medical School, Boston, MA, USA | Author | Revision of the manuscript for content, including medical writing for content, Major role in the acquisition of data, analysis or interpretation of data |
| Christina Jern, MD, PhD | Institute of Biomedicine, Department of Laboratory Medicine, the Sahlgrenska Academy, University of Gothenburg, Gothenburg, Sweden; Region Västra Götaland, Sahlgrenska University Hospital, Department of Clinical Genetics and Genomics, Gothenburg, Sweden | Author, Site PI | Revision of the manuscript for content, including medical writing for content, Major role in the acquisition of data, analysis or interpretation of data |
| Natalia S. Rost, MD, PhD | J. Philip Kistler Stroke Research Center, Massachusetts General Hospital, Harvard Medical School, Boston, MA, USA | Author, Site PI | Revision of the manuscript for content, including medical writing for content, Major role in the acquisition of data, analysis or interpretation of data |
| Arne G. Lindgren, MD, PhD | Section of Neurology, Skåne University Hospital, Lund, Sweden; Department of Clinical Sciences Lund, Neurology, Lund University, Lund, Sweden | Author, Site PI | Revision of the manuscript for content, including medical writing for content, Major role in the acquisition of data, analysis or interpretation of data |
| Anne-Katrin Giese, MD | J. Philip Kistler Stroke Research Center, Massachusetts General Hospital, Boston, MA, USA | Coinvestigator | Data acquisition |
| Oscar R. Benavente, MD | Department of Medicine, Division of Neurology, University of British Columbia, Vancouver, British Columbia, Canada | Coinvestigator, Site PI | Data acquisition |
| Tatjana Rundek, MD, PhD | Department of Neurology and Evelyn F. McKnight Brain Institute, Miller School of Medicine, University of Miami, Miami, FL, USA | Coinvestigator, Site PI | Data acquisition |
| Daniel Woo, MD | Department of Neurology and Rehabilitation Medicine, University of Cincinnati College of Medicine, Cincinnati, OH, and Department of Neurology, Jacobs School of Medicine and Biomedical Sciences, University of Buffalo, NY, USA | Coinvestigator, Site PI | Data acquisition |
| Reinhold Schmidt, MD | Department of Neurology, Clinical Division of Neurogeriatrics, Medical University Graz, Graz, Austria | Coinvestigator, Site PI | Data acquisition |
| Ramin Zand, MD | Department of Neurology, Geisinger, Danville, PA | Coinvestigator, Site PI | Data acquisition |
| Turgut Tatlisumak, MD, PhD | Department of Neurology, Helsinki University Hospital, Helsinkni, Finland, Department of Clinical Neuroscience, Institute of Neuroscience and Physiology, Sahlgrenska Academy at University of Gothenburg, Gothenburg, Sweden; Department of Neurology, Sahlgrenska University Hospital, Gothenburg, Sweden | Coinvestigator, Site PI | Data acquisition |
| Agnieszka Slowik, MD, PhD | Department of Neurology, Jagiellonian University Medical College, Krakow, Poland | Coinvestigator, Site PI | Data acquisition |
| James F. Meschia, MD | Department of Neurology, Mayo Clinic, Jacksonville, FL, USA | Coinvestigator, Site PI | Data acquisition |
| Jordi Jimenez-Conde, MD | Department of Neurology, Neurovascular Research Group (NEUVAS), IMIMHospital del Mar (Institut Hospital del Mar d’Investigacions M`ediques), Universitat Autonoma de Barcelona, Barcelona, Spain | Coinvestigator, Site PI | Data acquisition |
| Steven J Kittner, MD | Department of Neurology, University of Maryland School of Medicine and Veterans Affairs Maryland Health Care System, Baltimore, MD, USA | Coinvestigator, Site PI | Data acquisition |
| Bradford B. Worrall, MD, MSc | Departments of Neurology and Public Health Sciences, University of Virginia, Charlottesville, VA | Coinvestigator, Site PI | Data acquisition |
| Laura Heitsch, MD | Division of Emergency Medicine, Washington University School of Medicine, St Louis, MO; Department of Neurology, Washington University School of Medicine & Barnes- Jewish Hospital, St Louis, MO, USA | Coinvestigator, Site PI | Data acquisition |
| Pankaj Sharma, MD, PhD. FRCP | Institute of Cardiovascular Research, Royal Holloway University of London (ICR2UL), Egham, UK St Peter’s and Ashford Hospitals, UK | Coinvestigator, Site PI | Data acquisition |
| Arndt Rolfs, MD | Klinik und Poliklinik für Neurologie, Universitätsmedizin Rostock, Rostock, Germany | Coinvestigator, Site PI | Data acquisition |
| Robin Lemmens, MD | KU Leuven - University of Leuven, Department of Neurosciences, Experimental Neurology and Leuven Research Institute for Neuroscience and Disease (LIND), Leuven, Belgium; VIB, Vesalius Research Center, Laboratory of Neurobiology, University Hospitals Leuven, Department of Neurology, Leuven, Belgium | Coinvestigator, Site PI | Data acquisition |
| Vincent Thijs, PhD | Stroke Division, Florey Institute of Neuroscience and Mental Health, Heidelberg, Australia and Department of Neurology, Austin Health, Heidelberg, Australia | Coinvestigator, Site PI | Data acquisition |
